# The impact of studies with no events in both arms on meta-analysis of rare events: a simulation study using generalized linear mixed model

**DOI:** 10.1101/2021.08.23.21262461

**Authors:** Chang Xu, Lifeng Lin

## Abstract

**Objective:** The common approach to meta-analysis with double-zero studies is to remove such studies. Our previous work has confirmed that exclusion of these studies may impact the results. In this study, we undertook extensive simulations to investigate how the results of meta-analyses would be impacted in relation to the proportion of such studies.

**Methods:** Two standard generalized linear mixed models (GLMMs) were employed for the meta-analysis. The statistical properties of the two GLMMs were first examined in terms of percentage bias, mean squared error, and coverage. We then repeated all the meta-analyses after excluding double-zero studies. Direction of estimated effects and *p*-values for including against excluding double-zero studies were compared in nine ascending groups classified by the proportion of double-zero studies within a meta-analysis.

**Results:** Based on 50,000 simulated meta-analyses, the two GLMMs almost achieved unbiased estimation and reasonable coverage in most of the situations. When excluding double-zero studies, 0.00% to 4.47% of the meta-analyses changed the direction of effect size, and 0.61% to 8.78% changed direction of the significance of *p*-value. When the proportion of double-zero studies increased in a meta-analysis, the probability of the effect size changed the direction increased; when the proportion was about 40% to 60%, it has the largest impact on the change of *p*-values.

**Conclusion:** Double-zero studies can impact the results of meta-analysis and excluding them may be problematic. The impact of such studies on meta-analysis varies by the proportion of such studies within a meta-analysis.

## Introduction

Meta-analysis is a crucial statistical tool to synthesize information of similar studies and is commonly employed in medicine, education, sociology, psychology and related areas [1–4]. Standard meta-analytic methods based on asymptotic theory with a large sample assumption usually achieve unbiased estimation [5]. However, in the case of dichotomous outcomes, especially for rare outcomes (e.g., rare adverse events) or for outcomes where zero events were reported, the assumption no longer holds and may result in biased or even invalid estimation [6].

Studies with zero events are generally due to low incident rate, small sample size, or limited follow-up period. Dealing with studies of no events in both arms has been a tough problem because the relative estimates, such as odds ratio (OR), and the variance are hard to be defined [7–11]. A common approach for studies with no events in both arms is to apply a continuity correction of 0.5, while this method has been criticized for a number of drawbacks [12]. The more common approach is to remove such studies by treating them as non-informative [13]. However, some researchers argued that these studies may contain usable information for statistical inference and omitting them in analysis may lead to selection bias [14, 15].

Methodologists therefore developed several models to handle this issue. For example, Kuss et al. and Chu et al. proposed the beta-binomial model and their simulation verified the performance of this model [16, 17]. Liu et al. used the *p*-*function* model (within the exact framework [18]) based on Fisher’s *mid-p* correction to combine studies with no events [19]. Ren et al. compared the performance of standard methods (e.g., Peto, continuity correction), exact methods, and Bayesian methods, and recommended using the exact and Bayesian methods when there were studies with no events [20]. An alternative option was the generalized linear mixed model (GLMM) described by Stijnen et al. [21] and Simmonds and Higgins [22]. This model is a one-stage framework and allows studies with no events in both arms to “borrow strength” from studies with events without adding *a priori* assumption or *post hoc* correction [21–24]. Jackson et al. discussed six generalized linear mixed models for rare events and suggested that GLMMs performed better in the statistical inference than standard two-stage random-effects model [25].

Our previous work, by fitting a random-intercept GLMM model, has demonstrated that studies with no events contain important information and excluding them would have an impact on the results of meta-analysis [26]. We observed that the impact of studies with no events in both arms on meta-analysis may differ by the number of studies with no events in both arms within a meta-analysis. It is unclear whether the impact still exist with different number of studies with no events in both arms within a meta-analysis. In this article, we employed a large-scale simulation study to explore the impact of such studies on meta-analysis in terms of the proportion of them within a meta-analysis. For simplicity, we will refer “studies with no events in both arms” to as studies with no events in the following context.

## Methods

### Data generation

Our simulation is designed and reported according to the guidance from Pateras K et al, Burton A et al, and Morris TP et al [27–29]. The protocol of current simulation has been documented in our previous work [26]. We generated grouped data (2 × 2 table) in the simulation. The following fixed or random parameters were designed for current simulation: sample size of each trial (random), sample size ratio of two arms (random), number of studies within a meta-analysis (random), event risk in the control arm (fixed), true effect size (fixed), between-study variance (fixed). The true effect size and between-study variance were set to form different scenarios (see below). In order to generate a dataset that was aligned to the real-world settings, we performed a simulation with the parameter setting based on systematic reviews published from January 2003 to May 2018 in the Cochrane Database of Systematic Reviews (CDSR) [30, 31]. From the CDSR, we identified 442 meta-analyses (with data from 3,652 trials) which contained studies with no events (supplementary file 1) could be used for analysis [26].

The sample size information (the above 442 meta-analyses) was fitted into 71 commonly used distributions and the log-normal distribution fitted well in both the experimental and the control arms. Considering the potential correlations on the sample size of the two arms, we further analyzed the sample size ratio of them and utilized the ratio and the log-normal distribution of the control arm to get the sample size of the experimental arm. To be simple, here we took the uniform distribution of the first to third quartiles (0.84–2.04) of the sample size ratio (simply referred to as *ratio* henceforth) from the empirical data. Let *n*_l_ and *n*_2_ be the sample sizes of experimental and control arms; then, log *n*_2_ ~ *N* (3.3537, 0.9992), *n*_l_ = exp (log *n*_2_)\**ratio*, and *ratio* ~ uniform (0.84, 2.04).

The mean risk of the control arm from the 3,652 trials was 0.07; we however set it as 0.01 to improve the possibility for generating studies with zero events. This would allow us to get sufficient numbers of meta-analyses that contain studies with no events. The risk of the experimental arm was determined by effect size (i.e., OR) and the between-study variance (tau-square). To cover more scenarios, 5 monotonic increased values with equally-spaced ranges from small to large for effect size (1, 2, 3, 4, 5) and tau (0.2, 0.4, 0.6, 0.8, 1.0) were considered. Table 1 summarizes the simulation settings.

In the CDSR data, the study numbers in each meta-analysis ranged from 2 to 78, with the first to third quartiles as 4 and 10, respectively (Table S1, supplementary file 2). The study numbers was then randomly sampled from 4, 5, 6, 7, 8, 9 and 10. This setting also facilitated the calculation of the proportion (0% to 100%) of studies with no events in each meta-analysis. We generated 50,000 iterations for each scenario; among them, we removed those failed to contain studies with no events. We removed those meta-analyses with all studies as no events. In the GLMM model for meta-analysis, if the studies with the dependent variable was constant, we removed it owing to the basic assumption of likelihood-based methods for the parameter estimation [32, 33].

### Data analysis

The GLMMs were employed to combine data based on the link function “*logit*” under the “*binominal*” family. Here we considered two standard GLMMs for meta-analysis, i.e., the random slope GLMM and the random intercept and slope GLMM [25, 34]. Suppose there are *i* studies with *j* intervention status (1 for experimental and 0 for control). The random slope model employs random treatment effects *θ*_i_~*N*(*θ, τ*^2^) with fixed study effects (*γ*_*i*_) based on the hierarchical logistic regression within a frequentist framework: *logit*(*π*_*ij*_)= *γ*_i_ + *jθ*_*i*_. Here *θ* is log OR, *τ* is the between-study variance due to varies treatment effect, and π is the event risk. We call it the random slope GLMM since the constant term is generally excluded in the model [25, 34]. The random intercept and slope model employ both random study effects *γ*_*i*_ ~*N*(*γ, σ*^2^) and random treatment effects *θ*_*i*_ ~*N*(*θ, τ*^2^). The later accounts for the variance of study effects (*σ*^2^) across studies and is expected to be more conservative.

We first examined the statistical properties of GLMMs for combining studies with no events based on current parameter settings. The percentage bias, mean squared error (MSE), and coverage were utilized as indicators for the statistical properties of point and interval estimation. We further repeated the meta-analyses after excluding studies of no events to investigate the impact of studies of no events for meta-analysis. The direction of the estimated ORs and *p*-values for including and excluding studies with no events were subsequently compared. Paired meta-analyses that changed the direction of ORs and *p*-values were summarized. The threshold of the direction for OR was set as 1 and for *p*-value as 0.05 separately. For example, if an OR changed from >1 to <1 (or from <1 to >1), or a *p*-value changed from >0.05 to <0.05 (or from <0.05 to >0.05), then we considered that the direction was changed.

To investigate the potential impact of the number of studies with no events within each meta-analysis on the estimation, we stratified the whole samples into 9 ascending groups by the proportion of the number of studies with no events in a meta-analysis: 0.01%–10%, 10.01%–20%, 20.01%–30%, 30.01%–40%, 40.01%–50%, 50.01%–60%, 60.01%–70%, 70.01%–80%, and 80.01%–90%. The GLMMs were implemented by the *meglm* command in STATA/SE 14.0 (Stata, College Station, TX). The command for the random slope GLMM is: *meglm r study group || study: group, nocons family(binomial n) intme(lap)*. Also, the command for random intercept and slope GLMM is: *meglm r study group || study: group, family(binomial n) intme(lap)*. Where *r* is the number of cases and group is the intervention status. We used the Laplacian method for the estimation of the log likelihood; it is the default method for crossed random-effect model in Stata (version 14.0/SE, Stata, College Station, TX). The simulation code is presented in the supplementary file.

## Results

### Statistical properties of GLMMs

Figure 1 presents the percentage bias, MSE, and coverage of the two GLMMs under different scenarios. Our simulation suggested that the two GLMMs achieved almost unbiased estimation on the effect size in most of the situations when the between-study variance was not substantial (tau<0.8). We observed some extents of underestimation on effect size when true OR=1. Even when the between-study variance was large (tau=0.8 or 1.0), the percentage bias remained in a relative low level (1.89% to 35.2%). The MSE ranged from 0.78 to 1.23 for random slope GLMM and 1.03 to 1.32 for the random intercept and slope GLMM. A larger between-study variance or smaller effect size tended to lead to a larger MSE. Both GLMMs’ coverages were over the nominal level of 0.95 in all of the scenarios. The random intercept and slope GLMM showed lightly higher bias, MSE, and coverage than the random slope GLMM.

**Figure 1.**
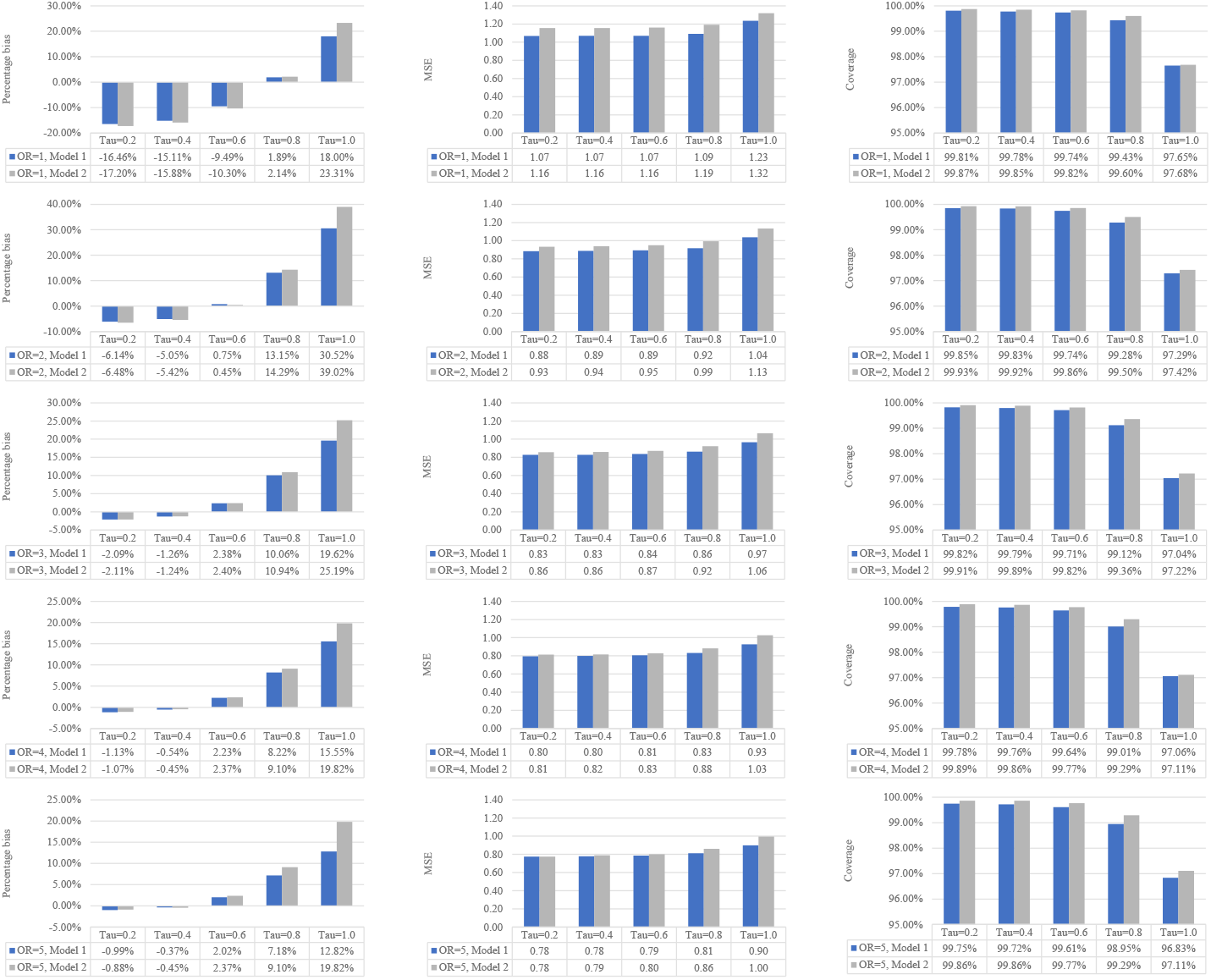
The statistical properties in terms of percentage bias, MSE, and coverage of the two GLMMs (Model 1: random slope GLMM. Model 2: random intercept and slope GLMM).

### The impact of studies with no events for meta-analysis

#### Effect size

Figure 2 presents the impact of studies with no events on the effect size for meta-analysis. In total, when excluding studies with no events, there were 0.0% (OR=5, tau=0.2) to 4.47% (OR=1, tau=1) of the meta-analyses with ORs changed the direction based on the random slope GLMM. A smaller proportion (0.0% to 2.47%) were observed in the random intercept and slope GLMM. The proportion varied in terms of the magnitude of effect size and between-study variance: those meta-analyses with larger effect size or smaller between-study variance tended to be more insensitive of the impact of studies with no events. There was almost no impact on the direction of OR when the true effect was large (e.g., OR=4 or 5).

**Figure 2.**
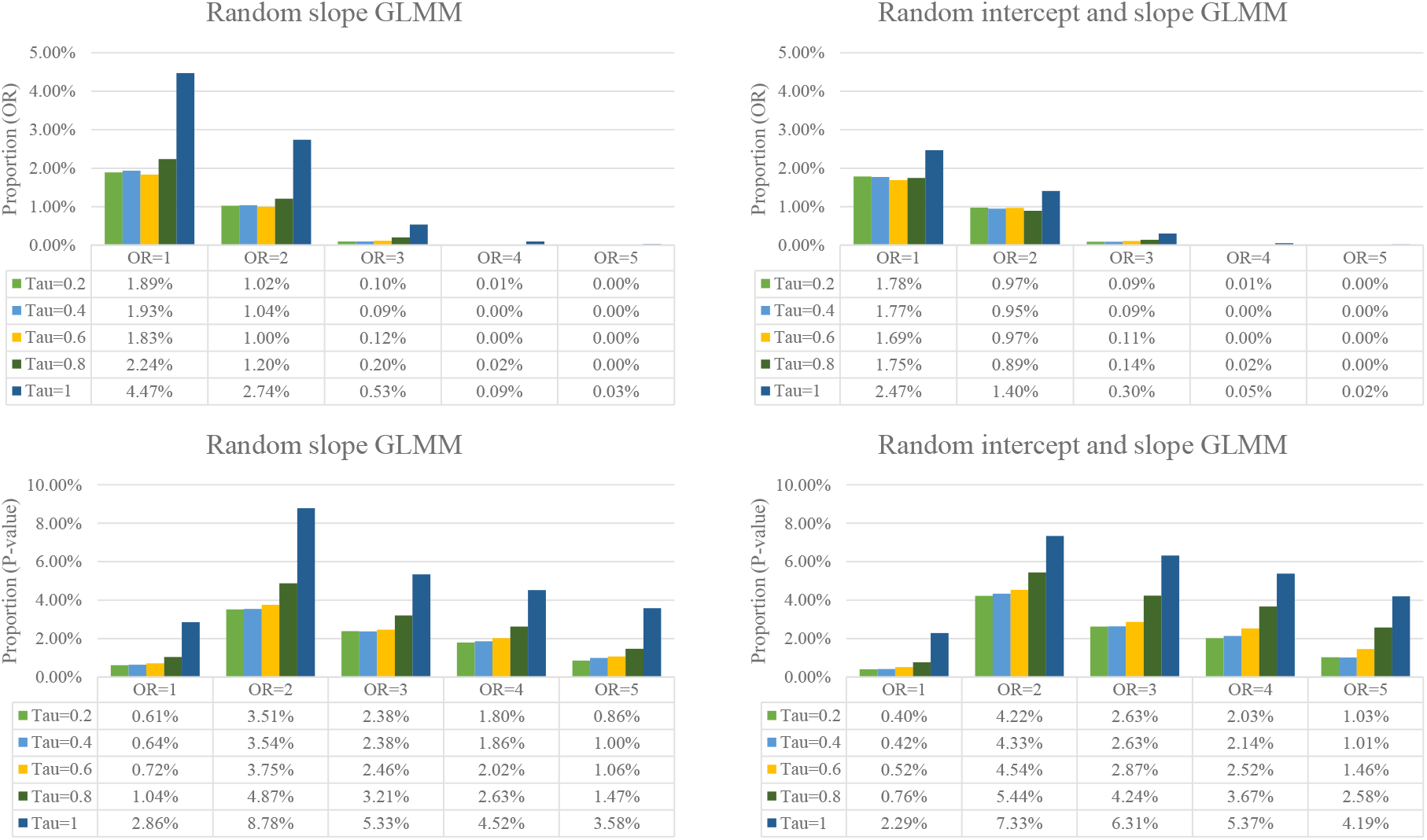
The impact of excluding studies with no events in both arms on the direction of effect size and significance of *p*-value.

#### P-value

In total, when excluding studies with no events, there were as many as 8.78% of the meta-analyses (OR=2, tau=1) with the *p*-value changed the direction of significance based on the random slope GLMM and 7.33% based on the random intercept and slope GLMM. Again, those meta-analyses with smaller between-study variance tended to be more insensitive of the impact of studies with no events. Interestingly, we observed that an OR=2 or 3 had the largest impact on the direction of *p*-value in both GLMMs (Figure 2).

### Number of studies with no events in a meta-analysis and the impact on the results

We stratified meta-analyses into 9 groups by the proportion (0.01%–90%) of the number of studies with no events. Table S2 (supplementary file 2) presents the number of meta-analyses in each group. Although we set a large-scale simulation, we noticed in some situations that there were insufficient sample sizes (less than 100) for the analysis where the proportion was high and the effect size was large (e.g., 80.01%–90%). The results of these scenarios should be treated with caution.

Our results suggested that, for effect size, when more studies with no events involved in a meta-analysis, the impact of excluding them would be larger (Figure 3 and Figure 4). In detail, when there was an increase in the proportion of studies with no events in a meta-analysis, there was a higher probability of the effect size (OR) changed the direction in all 25 scenarios. This was not the case for the direction of *p*-value on the significance (Figure 5 and Figure 6). For *p*-value, we observed that when the number of studies with no events accounted for 50% (centered around 40%–60%) in each meta-analysis, they would have the largest impact on the direction of *p*-value.

**Figure 3.**
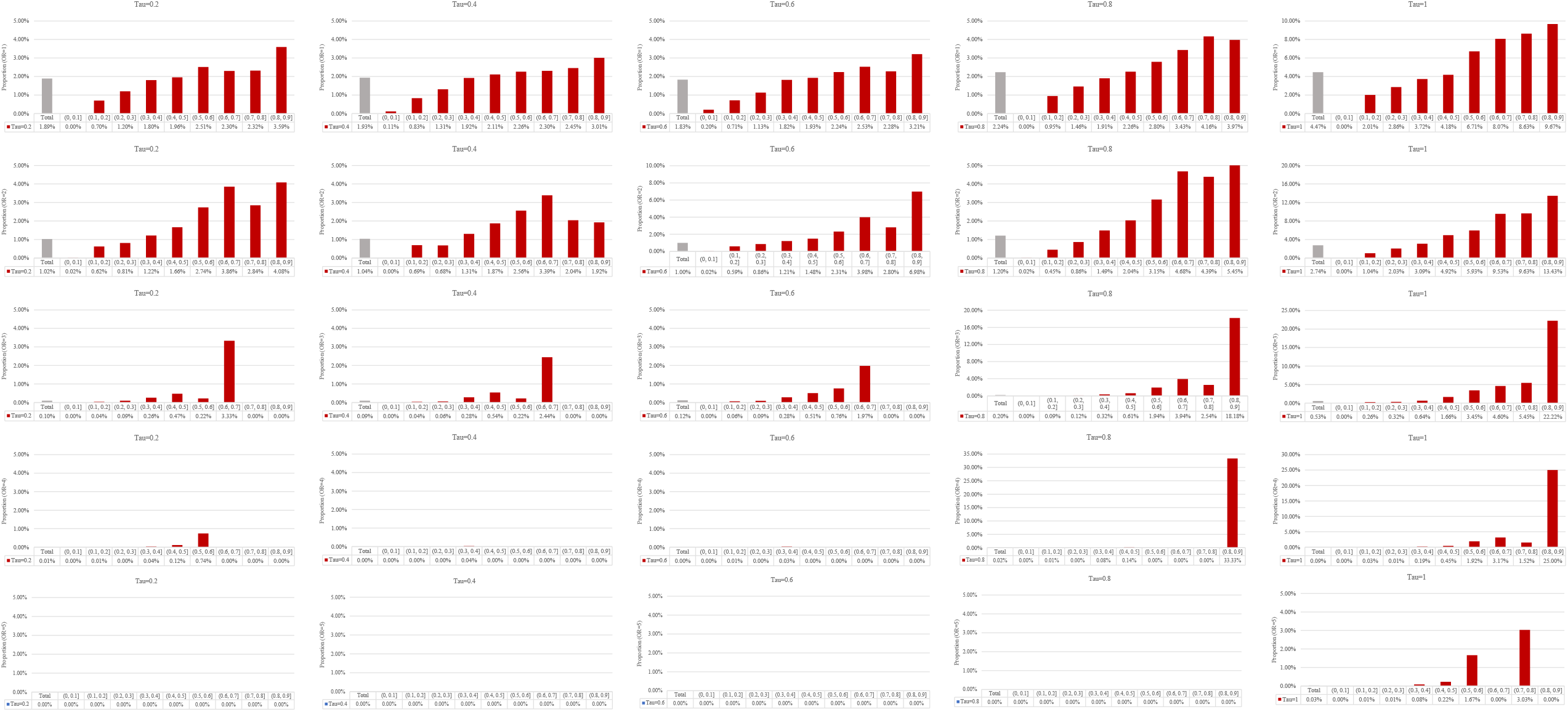
The proportion of OR changed the direction for including against excluding studies with no events based on the random slope GLMM.

**Figure 4.**
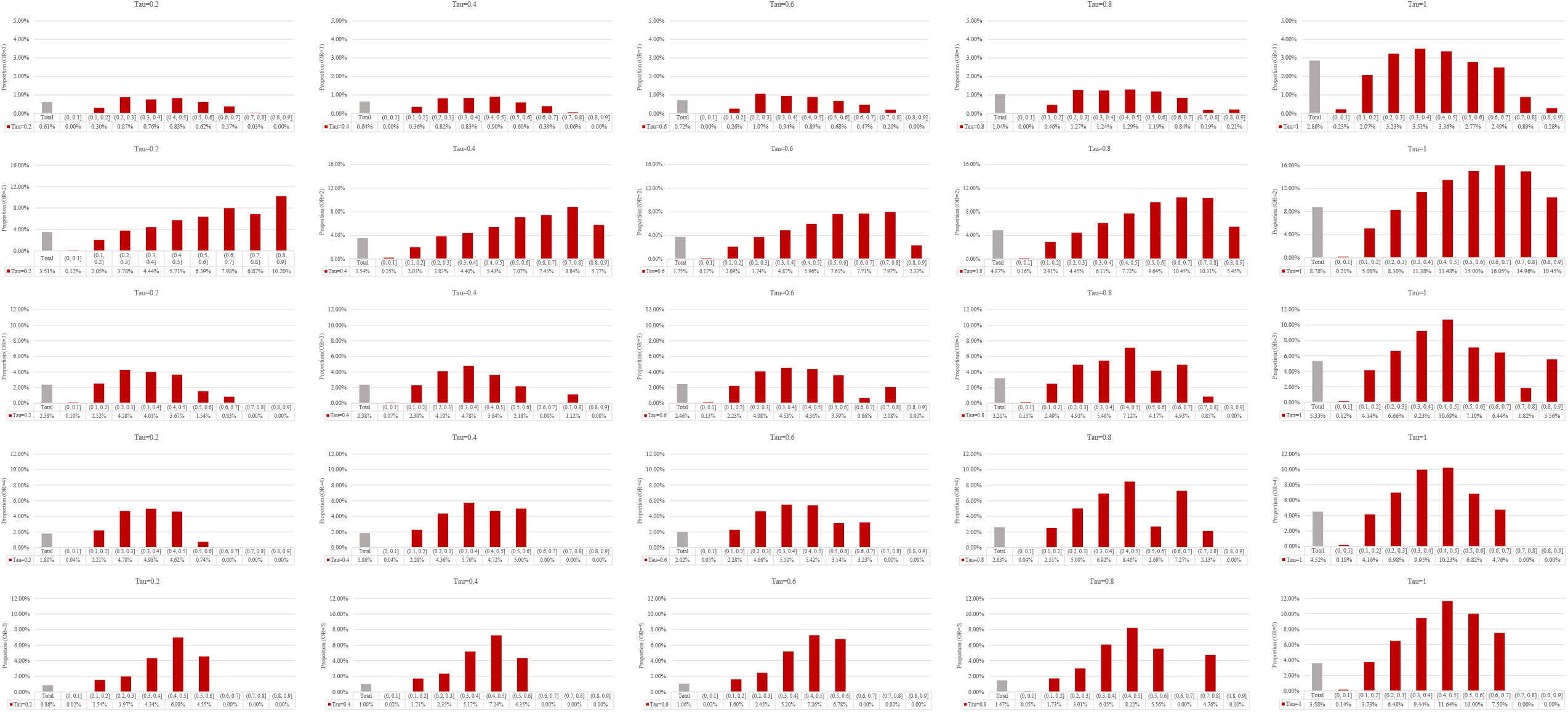
The proportion of OR changed the direction for including against excluding studies with no events based on the random intercept and slope GLMM.

**Figure 5.**
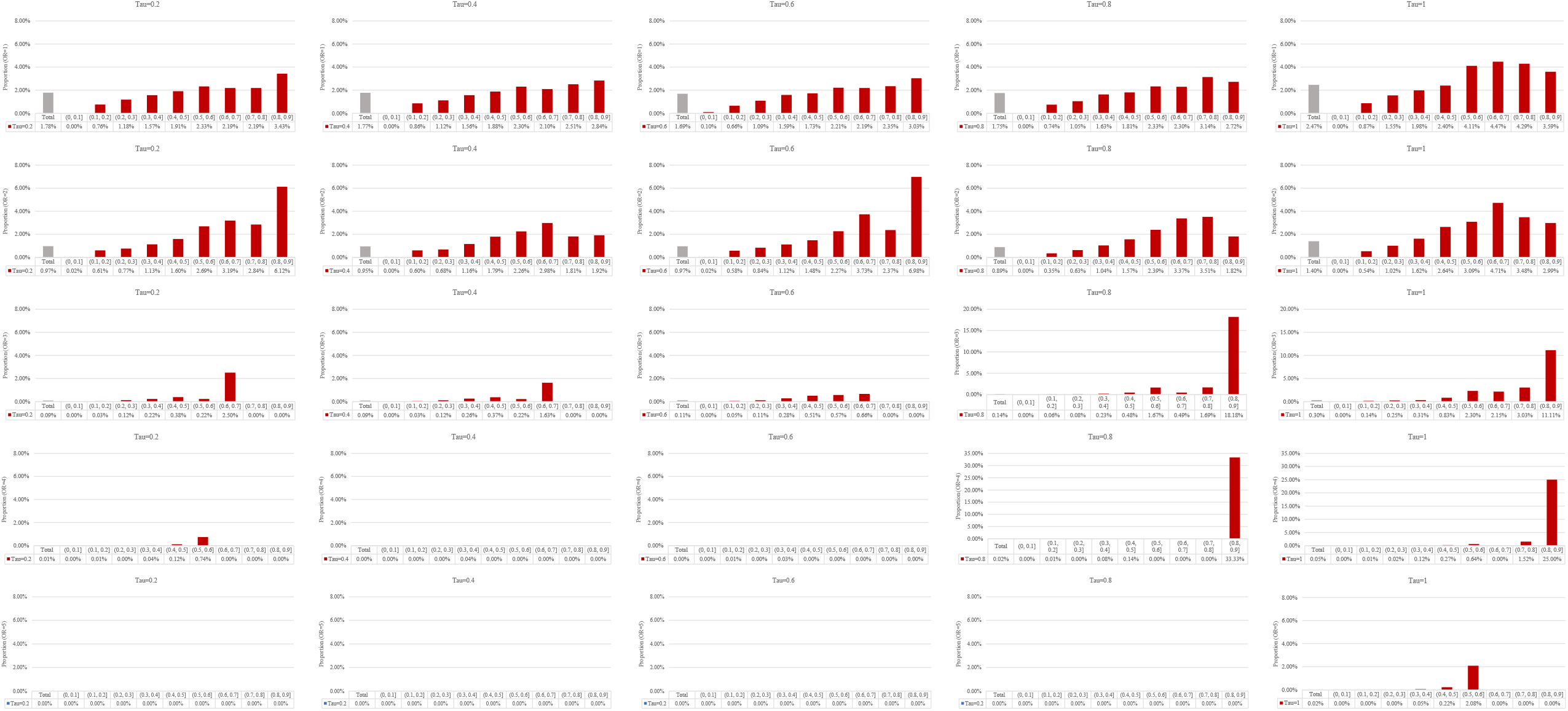
The proportion of *p*-value changed the direction of significance for including against excluding studies with no events based on the random slope GLMM.

**Figure 6.**
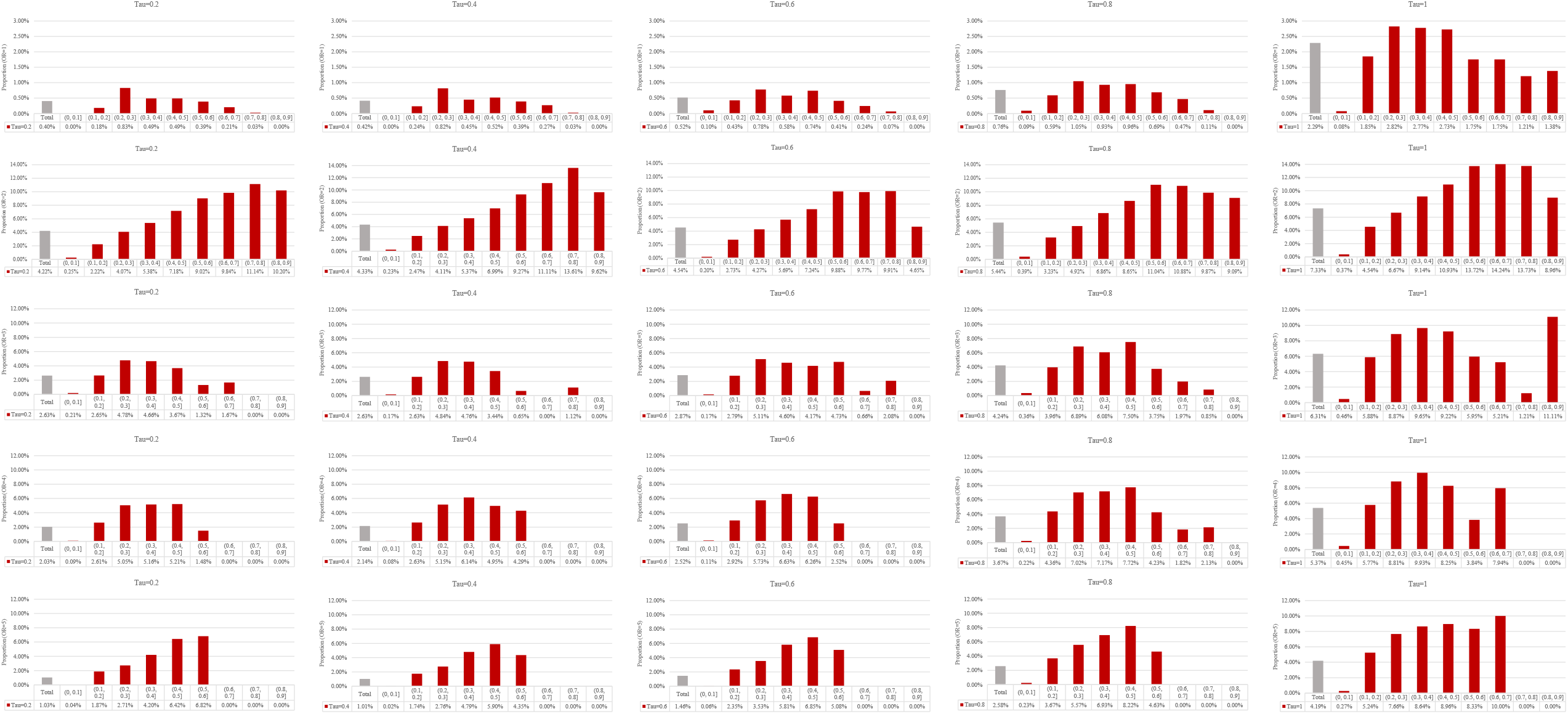
The proportion of *p*-value changed the direction of significance for including against excluding studies with no events based on the random intercept and slope GLMM.

As a result, when there was a low proportion of studies with no events in a meta-analysis (e.g., proportion≤10%), the impact of excluding studies with no events on the direction of OR and *p*-value was small (Figure 3–6). In this case, about 0.00% to 0.20% meta-analyses would change the direction on OR and 0.00% to 0.46% (0.25% in the random slope GLMM) would change the direction on *p-*value, regardless of the magnitude of effect size and between-study variance.

## Discussion

We conducted a large-scale simulation study to investigate the impact of studies with no events in both arms for the results of meta-analysis. Our findings verified that studies with no events in meta-analysis have considerable impact on the results that may change the conclusions. Although this impact was generally small on effect size, it was large on *p*-value. The impact depends on the proportion of the number of such studies within a meta-analysis: a higher proportion has a larger impact on the effect size, while a proportion of about 50% has the largest impact on the *p*-value. We further observed that for meta-analyses included studies with no events less than 10%, there would have the least impact on the direction of OR and *p-*value.

Studies with no events in both arms are commonly seen in meta-analyses of safety. Based on our ongoing research, there were 63.45% of the meta-analyses that with safety as exclusive outcome contains zero-events studies, and of which at least 50% contains studies with no events in both arms [35]. Such studies bring challenges for both statisticians and systematic review authors on how to deal with them. Currently, the routine approach is to simply remove them from the synthesis which simply treat them as “non-informative” [35]. Our study verified that such studies could not be treated as “non-informative” and the routine approach may be inappropriate that it’s time for a change.

In our simulation, when there was no true effect (OR=1) or the true effect was small (i.e. OR≤2), excluding studies with no events has considerable impact on the direction of OR. When the true effect was large (OR>3), studies with no events have limited impact on effect size. This can be expected because compared with a small effect that close to 1, it needs more “evidence” to push a large effect toward an opposite direction. However, these findings did not signify that studies with no events could be discarded when there was large effect. This is because excluding them is expected to change the between-study variance as well and further impacts the statistical inference.

We found that when studies with no events account for 50% of the numbers within a meta-analysis, it has the largest impact on the direction of *p*-value. This is because when the proportion is small, the weight of these studies is too weak to predominate the result; and when the proportion is large, these studies will however push the pooled effect to 1 as the expected OR of balanced studies with no events was 1, which make it hard to change the direction of *p*-value (when OR≈1, the *p*-value≈1 as well). Interestingly, we observed that moderate effect sizes (OR=2 or 3) have the largest impact on the direction of *p*-value. One possible explanation is that, for meta-analyses with OR=1 the *p*-value is large and converges to 1, and for meta-analyses with large effect (OR=4 or 5) the *p*-value is small and converges to 0; both are less sensitive to the impact of excluding studies with no events on the direction of *p*-values.

These findings from this simulation concurs with our previous study [26] where we observed that, based on empirical data modeled with the random intercept GLMM, when excluding studies with no events, for those meta-analyses changed the direction of *p*-values the proportion of studies with no events within a meta-analysis mainly clustered in 5%–30%. For meta-analyses that changed the direction of OR, the ORs mainly clustered in 0.84–1.04 [26]. The current simulation employed two random effect GLMMs to verify these findings. Based on our previous study and the current one, we found that the random effect GLMMs were less sensitive on the impact of excluding studies with no events for effect size than the random intercept GLMM, while comparable sensitive for *p*-values. Both these findings reinforce that studies with no events in both arms are not necessarily non-informative and should not be simply excluded from the synthesis [26, 36–37].

The current study verified the properties of GLMMs for pooling studies with no events in meta-analysis. One of the predominant advantages for GLMMs for meta-analysis is that they allow the estimation of the effect of studies with no events without requiring *a priori* assumption or *post hoc* correction. When the total events in both arms are sufficient (i.e., ≥10 [38]), the model would achieve a good estimation. In our previous simulation, we observed a very small percentage bias for GLMMs [26]. However, when the total events are small in either of the arms, the performance of GLMMs deteriorates due to the “small sample bias” of likelihood estimation. Researchers have proposed several solutions to this bias, for example, the penalized likelihood [39]. However, these methods have been verified to be not superior to the standard likelihood estimation [40] and the GLMMs would make no sense when the total events in either of the groups were zero. The beta-binomial model may be promising solutions for meta-analyses with insufficient number of total events [16,17, 41, 42]. Due to the complexity of such meta-analyses, further guidance on how to deal with studies with no events is needed.

## Conclusions

Based on our simulation, studies with no events contain information and have considerable impact on the results of a meta-analysis; the impact varies by the proportion of numbers of studies with no events. Excluding these studies is inappropriate and may lead to misleading conclusions and should be avoided in the future. The GLMMs could be a potential solution to combine studies with no events in meta-analysis.

## Data Availability

Data could be obtained through the following link.

https://www.researchgate.net/publication/350663359_The_impact_of_studies_with_no_events_in_both_arms_on_meta-analysis_of_rare_events_a_simulation_study_using_generalized_linear_mixed_model

## Availability of data and material

Available at: https://www.researchgate.net/publication/350663359_The_impact_of_studies_with_no_events_in_both_arms_on_meta-analysis_of_rare_events_a_simulation_study_using_generalized_linear_mixed_model

## Author’s contributions

CX conceived and designed the study, analyzed the data, and drafted the manuscript; CX and LL interpreted the results; LL provided careful comments on the statistical methods; all authors contributed careful edits for the manuscript. All authors approved the final version.

## Acknowledgement

None

