## Supplemental Tables for "The impact of studies with no events in both arms on meta-analysis of rare events: a simulation study using generalized linear mixed model"

Table S1. Baseline characteristics for 442 meta-analyses (3,652 trials).

| **Baseline characteristics** | **Meta-analyses with studies of no events** |
| --- | --- |
| **Number of studies within a meta-analysis** (ranges: 2 to 78), median (first to third quartile) | 6 (4 to 10) |
| <=6 | 244 (55.33%) |
| >6 | 197 (44.67%) |
| **Number of studies without events within a meta-analysis** (ranges: 1 to 33), median (first to third quartile) | 2 (1 to 3) |
| <=2 | 284 (64.40%) |
| >2 | 157 (35.60%) |
| *Proportion for studies without events to the all (ranges: 3.57% to 92.31%)* | 42.86% (25.00% to 57.14%) |
| **Sample size in each meta-analysis** (ranges: 40 to 18,034), median (first to third quartile) | 516 (276 to 1,206) |
| <=516 | 222 (50.23%) |
| >516 | 220 (49.77%) |
| **Sample size of studies without events in each study** (ranges: 2 to 1,397), median (first to third quartile) | 38 (22 to 71) |
| <=38 | 762 (20.83%) |
| >38 | 2,890 (79.17%) |
| *Proportion for studies without events to the all within a meta-analysis (ranges: 1.15% to 97.50%)* | 21.78% (10.10% to 39.45%) |

Note: The numbers in the right column were: median (first quartile to third quartile).

**The above table is from our published paper of the Table 1:**

Xu C, Li L, Lin L, et al. Exclusion of studies with no events in both arms in meta-analysis impacted the conclusions. Journal of Clinical Epidemiology 2020; 123:91-99.

**Table S2. Number of simulated meta-analyses within each group stratified by the proportion of studies with no events within a meta-analysis.**

| **Proportion of studies with no events within MA** | **OR=1** | | | | | **OR=2** | | | | | **OR=3** | | | | | **OR=4** | | | | | **OR=5** | | | | |
| --- | --- | --- | --- | --- | --- | --- | --- | --- | --- | --- | --- | --- | --- | --- | --- | --- | --- | --- | --- | --- | --- | --- | --- | --- | --- |
|  | Tau=0.2 | Tau=0.4 | Tau=0.6 | Tau=0.8 | Tau=1.0 | Tau=0.2 | Tau=0.4 | Tau=0.6 | Tau=0.8 | Tau=1.0 | Tau=0.2 | Tau=0.4 | Tau=0.6 | Tau=0.8 | Tau=1.0 | Tau=0.2 | Tau=0.4 | Tau=0.6 | Tau=0.8 | Tau=1.0 | Tau=0.2 | Tau=0.4 | Tau=0.6 | Tau=0.8 | Tau=1.0 |
| Group 1 (0.01%~10%) | 872 | 894 | 978 | 1,107 | 1,323 | 5,680 | 5,550 | 5,444 | 5,141 | 4,838 | 12,911 | 12,710 | 12,138 | 10,737 | 9,050 | 19,898 | 19,670 | 18,512 | 16,193 | 13,172 | 25,566 | 25,236 | 23,791 | 20,920 | 16,817 |
| Group 2 (10.01%~20%) | 3,288 | 3,366 | 3,508 | 3,901 | 4,487 | 11,239 | 11,240 | 10,980 | 10,711 | 10,219 | 15,449 | 15,423 | 15,134 | 14,614 | 13,822 | 15,548 | 15,536 | 15,714 | 15,796 | 15,547 | 13,816 | 13,963 | 14,571 | 15,274 | 15,850 |
| Group 3 (20.01%~30%) | 4,584 | 4,651 | 4,878 | 5,340 | 5,880 | 9,522 | 9,567 | 9,578 | 9,540 | 9,447 | 8,577 | 8,680 | 8,922 | 9,319 | 9,706 | 6,123 | 6,235 | 6,616 | 7,537 | 8,504 | 4,056 | 4,131 | 4,616 | 5,743 | 7,258 |
| Group 4 (30.01%~40%) | 8,598 | 8,633 | 8,790 | 9,027 | 9,586 | 9,458 | 9,410 | 9,444 | 9,539 | 9,676 | 5,384 | 5,421 | 5,784 | 6,481 | 7,316 | 2,731 | 2,849 | 3,198 | 3,961 | 5,205 | 1,476 | 1,566 | 1,789 | 2,495 | 3,727 |
| Group 5 (40.01%~50%) | 11,242 | 11,240 | 11,245 | 11,296 | 11,007 | 6,202 | 6,299 | 6,475 | 6,681 | 7,054 | 2,341 | 2,416 | 2,566 | 3,120 | 3,862 | 845 | 848 | 1,071 | 1,477 | 2,229 | 358 | 373 | 482 | 754 | 1,340 |
| Group 6 (50.01%~60%) | 6,959 | 6,856 | 6,780 | 6,403 | 5,988 | 1,973 | 1,995 | 2,075 | 2,219 | 2,427 | 454 | 459 | 529 | 720 | 1,042 | 135 | 140 | 159 | 260 | 469 | 44 | 46 | 59 | 108 | 240 |
| Group 7 (60.01%~70%) | 4,832 | 4,819 | 4,513 | 4,048 | 3,533 | 752 | 738 | 778 | 919 | 997 | 120 | 123 | 152 | 203 | 326 | 27 | 26 | 31 | 55 | 126 | 4 | 5 | 9 | 15 | 40 |
| Group 8 (70.01%~80%) | 3,239 | 3,181 | 2,979 | 2,642 | 2,144 | 422 | 441 | 464 | 456 | 488 | 82 | 89 | 96 | 118 | 165 | 22 | 24 | 27 | 47 | 66 | 11 | 11 | 14 | 21 | 33 |
| Group 9 (80.01%~90%) | 612 | 599 | 561 | 478 | 362 | 49 | 52 | 43 | 55 | 67 | 8 | 5 | 4 | 11 | 18 | 1 | 1 | 0 | 3 | 8 | 0 | 0 | 0 | 0 | 2 |

Highlighted cells were those with insufficient number of meta-analyses.
